# Safety of Continuous Erector Spinae Catheters in Chest Trauma: A Retrospective Cohort Study

**DOI:** 10.1101/2021.05.06.21256789

**Authors:** L.D White, B. Riley, K. Davis, C. Thang, A. Mitchell, C. Abi-fares, W. Basson, C. Anstey

## Abstract

The erector spinae block is an efficacious analgesic option for the management of rib fracture related pain. Despite there being minimal published data specifically addressing the safety profile of this block, many societies have made statements regarding its safety and its use as an alternative to traditional regional anaesthesia techniques in patients at risk of complications. The primary aim of this study was to characterise the safety profile of erector spinae plane block catheters by determining the incidence of early complications. The secondary aims of this study was to characterise the incidence of late adverse events, as well as, the erector spinae plane block catheter failure rate. We analysed electronic medical record data of patients who had an erector spinae plane block catheter inserted for the management of rib fractures between November 2017 to September 2020. To assess early adverse events data collection included hypotension, hypoxaemia, local anaesthetic systemic toxicity and pneumothorax thought to be associated with erector spinae plane block catheter insertion. Late complications included catheter site infection and catheter site haematoma. Two hundred and twenty four patients received a total of 244 continuous erector spinae catheters during the study period. Following the insertion of the erector spinae there were no immediate complications such as hypotension, hypoxia, local anaesthetic toxicity or pneumothorax. Of all blocks inserted 7.7% were removed due to catheter failure (OR = 8.4 per 100 catheters; 95%CI = 5.1 to 13.9 per 100 catheters). This resulted in a failure rate of 1.9 per 1000 catheter days (95%CI = 1.1 to 6.7 catheter days). Late complications included two erythematous catheter sites and two small haematomas not requiring intervention. The odds of a minor late complication was 16.7 per 1,000 catheters (95%CI = 6.1 to 45.5 per 1,000 catheters). In conclusion, this study supports the statements made by regional anaesthesia societies regarding the safety of the erector spinae plane block. Based on the results presented in this population of trauma patients, the erector spinae plane block catheter is a low risk analgesic technique which may be performed in the presence of abnormal coagulation status or systemic infection.

## Introduction

Rib fractures are common in patients presenting to the emergency department following blunt trauma. There is a high likelihood of morbidity, short and long-term disability and mortality associated with rib fractures [1-3]. Thoracic complications are the leading cause of death in this patient population. These complications include pneumonia, atelectasis and pulmonary emboli, arising as a result of hypoventilation secondary to pain and altered respiratory mechanics with complicating damage to underlying lung parenchyma [4-6]. As such, adequate pain control is paramount in decreasing the risk of death [7].

The pain resulting from rib fractures is notoriously difficult to manage and peaks in severity approximately five days post injury. In the past, opioids have been the mainstay of treatment however they have significant side effects including respiratory depression and sedation [5]. The gold standard for analgesia includes the early use of a regional anaesthesia catheter techniques such as the thoracic epidural and the paravertebral block [8-10]. However, given the complexity of both the injuries sustained during the inciting event and the patient’s comorbidities, there are often contraindications to the insertion of these blocks [11]. Furthermore, this patient population is at high risk of requiring complex anticoagulation or developing infections which would either preclude the insertion of or necessitate the early removal of an epidural or paravertebral catheter shortly after insertion.

Recently there has been a surge in case reports, case series and retrospective studies celebrating the success of more superficial chest wall blocks such as the erector spinae block [12-16]. The use of an erector spinae plane block has been reported to provide good analgesia with minimal complications [17-19]. The theory behind using these blocks is that they are in superficial, compressible locations and based on a risk benefit analysis could be utilised in patients with active infection or anticoagulation [10,17]. This opinion has been backed by the American Society for Regional Anesthesia(17). However, there have been no studies that have been performed with the primary aim of assessing the safety profile of the erector spinae plane block.

The primary aim of this study was to characterise the safety profile of erector spinae plane block catheters by determining the incidence of early and late complications. The secondary aim of this study was to characterise the associated erector spinae plane block catheter failure rate.

## Methods

This retrospective study was approved by The Prince Charles Hospital Human Research Ethics Committee and the Sunshine Coast Hospital and Health Service (HREC: LNR/2018/QPCH/45155). We analysed electronic medical record data of patients who had an Erector Spinae plane catheter inserted for the management of rib fractures between November 2017 to September 2020. Data from other superficial chest wall block catheters such as the serratus anterior block were excluded from this study. All data was retrieved and compiled into a standardised Microsoft Excel Spreadsheet (Microsoft Corp, Redmond, WA, USA; 2016). Demographic data included: age, medical comorbidities, mechanism of injury, injuries sustained and hospital length of stay.

All erector spinae plane blocks were performed by consultant anaesthetists or anaesthesia registrars. The clinicians performing this block insertion had a range of experience performing an erector spinae plane block. All novice clinicians were directly supervised by anaesthetists with significant experience in the insertion of erector spinae plane block catheters. All patients had erector spinae catheters sited under ultrasound guidance using a high frequency linear-array transducer (12-15 MHz, X-Porte, Sonosite, Bothell, MA) using a variety of Toohey needle sets. In the majority of cases, saline was used for the initial hydrodissection and the initial local anaesthetic loading dose was bolused via the catheter. The most common local anaesthetic loading doses at our institution was 15-30 mLs of 0.2% to 0.375% ropivacaine followed by an intermittent bolus one to three hourly of 0.2% ropivacaine (10-20mLs). When bilateral fractures were present, an erector spinae plane block was performed on both sides. Following insertion, the catheters were secured with glue, steri-strips and a clear adhesive dressing. The insertion sites were checked daily by the acute pain service until the day of removal and in most cases, included the day after catheter removal.

The primary outcome of interest was immediate adverse events associated with erector spinae plane catheters. The electronic medical records for each patient were manually searched for immediate adverse events as a result of the catheter insertion. This included hypotension during insertion, hypoxaemia during insertion, symptoms of Local Anaesthetic Systemic Toxicity and the diagnosis of a new pneumothorax thought to be associated with erector spinae plane catheter insertion. Secondary outcomes included late complications and erector spinae catheter failure rate. Late complications included either catheter site infection and catheter site haematoma. In the event of catheter site infection or catheter site haematoma, the management of these complications was assessed and treated accordingly.

The secondary outcome of this study was erector spinae plane catheter failure rate. To assess this outcome, data collected included duration that each catheter was left in situ and the reason for catheters removal.

Data analysis was performed using Microsoft Excel 2016 (Microsoft Corp, Redmond, WA, USA) and STATA_TM_ (ver 16.0) (StataCorp, College Station, Texas, USA). Data was presented as mean (standard deviation (SD)) and frequency (%) as required. Primary and secondary outcomes were presented as the number of events, percentage of events and the Odds Ratio (OR) with 95% Confidence Intervals (95%CI) per 100 or 1,000 catheters. Failure rate was calculated as the number of failed catheters per catheter day using the number of failed catheters and the time that the catheters were left in situ. Results were reported as the mean with 95%CI.

## Results

Two hundred and twenty four patients received a total of 244 continuous erector spinae catheters during the study period (Table 1). The mean age for these patients was 61.8 (16.2) years of age with an average of 6.1 (2.7) fractured ribs. The number of ribs fractured ranged one to seventeen. Of these 86.2% were unilateral, 32.6% had a flail chest, 56.7% had either a pneumothorax or a haemothorax, 32.1% had a pulmonary contusion and 28.6% required a chest drain. Comorbidities in this patient cohort included obstructive or restrictive lung disease (20.1%), ischaemic heart disease (16.5%), obstructive sleep apnoea (5.4%), liver disease (6.7%), renal disease (8.9%) and a smoking history (34.4%). The average duration (standard deviation) of hospital stay amongst the study population was 10.3(9.9) days. Prior to performing the erector spinae block, pre-existing complications and adverse physiological states included pneumonia (4.9%), therapeutic anticoagulation or antiplatelet (20.1%), systemic infection (6.7%) and deranged coagulation studies or low platelets (8.5%) (Table 1).

**Table 1:**
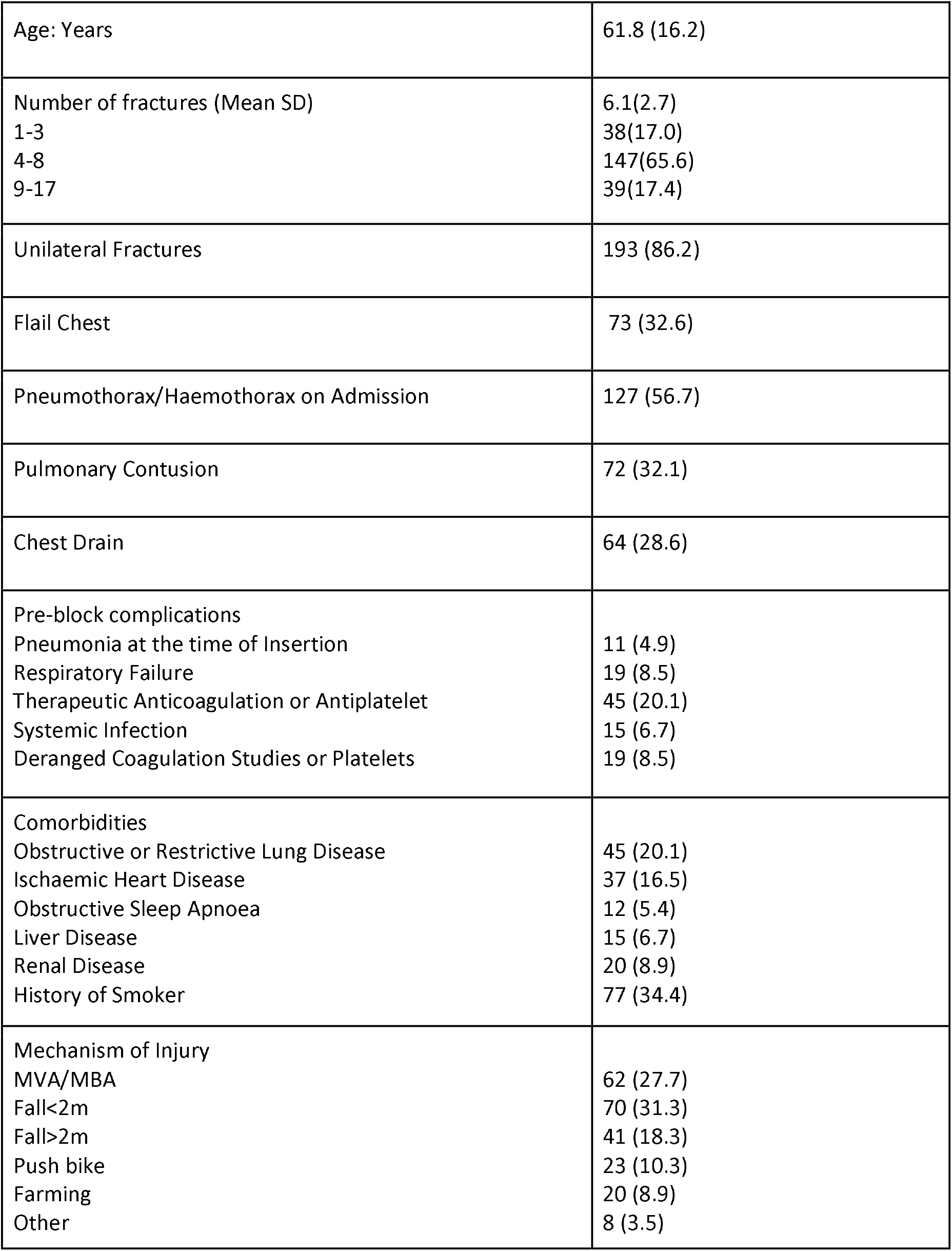
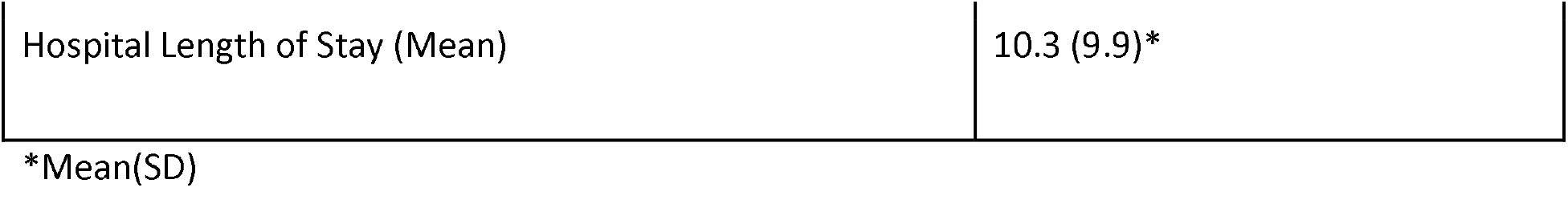
Patient characteristics

Following the performance of the erector spinae plane block there were no immediate complications as a result of catheter insertions such as hypotension, hypoxia, local anaesthetic systemic toxicity or pneumothorax (Table 2). The average duration of catheter dwell site that the catheter was left in situ was 4.1(1.5) days (Range of 1 to 8 days). Of all blocks inserted 7.7% were removed due to catheter failure (OR = 8.4 per 100 catheters; 95%CI = 5.1 to 13.9 per 100 catheters). This resulted in a failure rate of 1.9 per 1000 catheter days (95%CI = 1.1 to 6.7 catheter days). Late complications included two erythematous catheter sites and two small haematomas. Neither required an intervention. The odds of a late complication was 16.7 per 1,000 catheters (95%CI = 6.1 to 45.5 per 1,000 catheters). For catheters displaying both erythema and/or haematoma the removal rates was 8.3 per 1,000 catheters (95%CI = 2.0 to 33.6 per 1,000 catheters). None of the patients with catheters removed for erythema or haematoma required intervention.

**Table 2:** Safety Outcomes for the Erector Spinae Plane Block Catheter.

|  |  |
| --- | --- |
| Immediate/Insertion Related Complications | 0(0,0) |
| - Hypotension | 0(0.0) |
| - Hypoxaemia | 0(0.0) |
| - Local Anaesthetic Toxicity | 0(0.0) |
| - Pneumothorax | 0(0.0) |
| Number of Days Catheter In Situ | 4.1(1.5)* |
| Catheter Failure | 19(7.7) |
| Catheter site infection | 2(0.8) |
| - Erythema No Intervention | 2(0.8) |
| - Requiring Intervention | 0(0.0) |
| Catheter site haematoma | 2(0.8) |
| - Intervention Not Required | 2(0.8) |
| - Intervention Required | 0(0.0) |

\*Mean(SD)

## Discussion

This study investigated the incidence of adverse events related to the insertion of an erector spinae plane catheter for the management of rib fractures. Our results provide the incidence of immediate complications such as; hypotension; hypoxia; local anaesthetic systemic toxicity; pneumothorax, as well as, catheter failure rates, the incidence of catheter site infection and haematoma from 244 catheters over a four-year study period. Although the safety profile of an erector spinae plane block has not previously been studied, these results are consistent with expert opinion and studies primarily assessing analgesic efficacy [17,19]. This is an important addition to current literature, which supports the use of this efficacious regional anaesthetic technique for patients precluded from having a paravertebral block or thoracic epidural.

Over a quarter of patients included in the present study had contraindications to the performance of a thoracic epidural or paravertebral block. These contraindications included a significant risk of infection or bleeding related complications. However, no significant complications occurred as a result of erector spinae plane catheter insertion. On a thorough, manual review of patient electronic medical records, only two minor haematomas and two erythematous catheter sites were identified. Most importantly, none of these complications required an intervention to assist with their resolution. Up until now, the justification for the erector spinae plane block being safe, particularly in the context of anticoagulation, has been based on the compressibility and low consequences of bleeding at the site [10,20]. Therefore, the results of this study provides further justification for the classification of the erector spinae plane block as a superficial, compressible, low risk block.

Currently, our understanding of the exact analgesic mechanism of the erector spinae plane block is lacking [21-22]. One proposed mechanism is through the spread of local anaesthetic to the paravertebral and epidural space [21]. Local anaesthetic injection into the paravertebral space can be associated with significant haemodynamic events related to it’s sympatholytic activity [23]. An additional proposed benefit of the erector spinae plane block is its lack of haemodynamic effects. For this reason we assessed patient notes to confirm this proposed benefit. The results of the present study found no documented evidence of significant haemodynamic effects immediately following erector spinae catheter loading. Even though no haemodynamic events were documented, this does not refute or change our understanding of the proposed mechanism of analgesia. The incidence of clinically significant hypotension following the performance of a paravertebral block is approximately 4.9% [24].

An explanation of paravertebral blockade related hypotension is the effect on the sympathetic ganglia and spread into the epidural space, both of which are reduced with lower volumes and concentrations of local anaesthetic [23]. The slow, low volume spread of local anaesthetic from the erector spinae plane to the paravertebral and epidural spaces is likely why we did not find any hypotensive episodes in this study.

The decision to provide regional anaesthetic techniques for intraoperative and postoperative analgesia is based on the both the real, and perceived risk benefit ratio. The decision to provide regional anaesthetic techniques for intra and post operative analgesia is based on a patient centred approach including a weighted risk benefit analysis. Current evidence supports the use of regional anaesthetic techniques in patients at risk of complications from systemic analgesia. As the risk of systemic analgesia lessens the decision to provide regional anaesthesia becomes less clear, as evidenced by the controversies among certain patient groups. The decision to provide regional anaesthesia becomes less clear as patient risk related to systemic analgesia lessens. An example of this is the decision to perform a paravertebral block in elective cosmetic breast surgery [25]. While significant complications from this block is infrequent, some clinicians may still find the risk too high to justify it as part of their routine practice [23]. The erector spinae block is a valuable alternative to the paravertebral block, shown to have good efficacy in breast surgery [26-27]. The patient population included in this study was of higher risk of complications when compared to the typical elective surgery patient population, due to the nature of traumatic injuries sustained. Therefore, we believe this study fills an important gap for elective surgical patients that could be explored further. The erector spinae block is a low risk analgesic option in patients with a low risk benefit ratio relating to the decision to perform regional anaesthesia.

Similar to the paucity of evidence surrounding complication rates, there is no literature surrounding the longevity of a working erector spinae catheter. This is important as erector spinae catheters are often inserted in the first one or two days of admission. However, the pain related to acute rib fractures peaks at day five [28]. For this reason, a functioning erector spinae plane catheter needs to remain in situ long enough to successfully cover the acute pain phase, without an increased risk of complications. Our results showed a low failure rate of 7.7%. On average catheters remained in situ for 4.1 days to a maximum of eight days. The insertion of erector spinae catheters were performed by clinicians with varying skill levels in the insertion of an erector spinae block, ranging from novice to expert. Therefore, this enhances the external validity of the results presented in this study, transferable to clinicians of all skill levels.

There were many limitations to this retrospective study. The incidence of complications was limited by documentation in the electronic medical records. Every effort was made to identify adverse events with authors reading full patient files including the insertion notes, observation records and all medical reviews from the in patient stay. The follow up of the patient was limited to their inpatient stay. Therefore, late infective complications identified following patient discharge may have been missed. Additionally, our data on clinician experience in the performance of an erector spinae plane block was lacking. Therefore we were unable to make any comment on catheter failure rates in relation to clinician skill level.

In conclusion, this study supports the statements made by regional anaesthesia societies regarding the safety of the erector spinae plane block. In our study of 244 erector spinae plane catheters, there were no immediate or significant late complications. The catheter failure rate in this study was low (1.9 per 1000 catheter days) with a mean duration of time in situ of 4.1 days. Based on the results presented in this population of trauma patients, the erector spinae plane block catheter is a low risk analgesic technique which may be performed in the presence of abnormal coagulation status or systemic infection.

